# All Models Are Useful: Bayesian Ensembling for Robust High Resolution COVID-19 Forecasting

**DOI:** 10.1101/2021.03.12.21253495

**Authors:** Aniruddha Adiga, Lijing Wang, Benjamin Hurt, Akhil Peddireddy, Przemyslaw Porebski, Srinivasan Venkatramanan, Bryan Lewis, Madhav Marathe

## Abstract

Timely, high-resolution forecasts of infectious disease incidence are useful for policy makers in deciding intervention measures and estimating healthcare resource burden. In this paper, we consider the task of forecasting COVID-19 confirmed cases at the county level for the United States. Although multiple methods have been explored for this task, their performance has varied across space and time due to noisy data and the inherent dynamic nature of the pandemic. We present a forecasting pipeline which incorporates probabilistic forecasts from multiple statistical, machine learning and mechanistic methods through a Bayesian ensembling scheme, and has been operational for nearly 6 months serving local, state and federal policymakers in the United States. While showing that the Bayesian ensemble is at least as good as the individual methods, we also show that each individual method contributes significantly for different spatial regions and time points. We compare our model’s performance with other similar models being integrated into CDC-initiated COVID-19 Forecast Hub, and show better performance at longer forecast horizons. Finally, we also describe how such forecasts are used to increase lead time for training mechanistic scenario projections. Our work demonstrates that such a real-time high resolution forecasting pipeline can be developed by integrating multiple methods within a performance-based ensemble to support pandemic response.

**ACM Reference Format:** Aniruddha Adiga, Lijing Wang, Benjamin Hurt, Akhil Peddireddy, Przemys-law Porebski,, Srinivasan Venkatramanan, Bryan Lewis, Madhav Marathe. 2021. All Models Are Useful: Bayesian Ensembling for Robust High Resolution COVID-19 Forecasting. In *Proceedings of ACM Conference (Conference’17)*. ACM, New York, NY, USA, 9 pages. https://doi.org/10.1145/nnnnnnn.nnnnnnn

## 1 INTRODUCTION

COVID-19 pandemic has been the greatest public health challenge facing humanity in over a century with more than 100 million confirmed cases worldwide, leading to nearly 2.3 million recorded deaths. United States has been the most affected country with approximately 25% and 20% of the reported cases and deaths respectively. While early response was marred by lack of adequate testing and medical resources, subsequent surges can be attributed to uncoordinated response across spatial scales. Although multiple crowd-sourced^1^ and academic^2^ efforts led to databases on high resolution disease surveillance, actionable forecasts for early indicators were difficult to integrate into policy response.

For instance, in April 2020, the Centers for Disease Control and Prevention (CDC) in collaboration with academic partners initiated the COVID-19 Forecast Hub (hereon referred to as *The Hub*), a consortium of modeling teams to coordinate the forecasting efforts^3^. Due to lack of reliable data on other outcomes and at higher resolutions, this early effort was limited to probabilistic forecasts of deaths at the national and state level. Multiple classes of statistical and mechanistic models were employed by individual groups[31] and the forecasts were combined through a naive equally-weighted ensemble of eligible methods. Unlike seasonal influenza forecasting where statistical methods have been shown to be better overall[32], due to lack of training data and the need for integrating various disease parameter estimates, it was noted that mechanistic models such as the Susceptible-Infected-Recovered (SIR) and its variants proved to be more useful for COVID-19 forecasting. However, due to deaths being a lagging indicator of disease activity, it proved difficult to use for guiding policy interventions in real-time. Subsequently this effort was expanded in July 2020 to include incident case forecasts at the county level.

Forecasting disease incidence at finer spatial resolution is impacted by multiple factors such as: (a) higher noise partly due to lower population counts, (b) lack of exogenous observables such as mobility or testing rates at equivalent resolution, and (c) greater level of connectivity across regions leading to interdependence. Further, the COVID-19 experience across the United States has been quite heterogeneous spatially and temporally, making it difficult to obtain sufficient training data and reasonably track the different phases of the pandemic. Other aspects such as reporting errors, back-filled cases may lead to uncharacteristic spikes not necessarily reflecting the state of the pandemic. Finally, for geographically and demographically heterogeneous states like California and Virginia, it is difficult to *understand* the state level epidemic trajectories without considering the evolution at finer resolution, such as the county level. Thus, beyond aiding targeted interventions and resource demand estimation, forecasting at the county level for United States can help improve the overall forecasting quality and robustness at the state and national level. It is to be noted that of the nearly 70 teams contributing to *The Hub*, only 20% of them produce county level forecasts, many of them starting in late 2020. Further, not every team provides probabilistic forecasts due to the aforementioned challenges complicating uncertainty quantification. Since it is well accepted that ensemble forecasts tend to have better performance compared to individual forecasts [32, 37, 43], *The Hub* employs a non-performance-based ensemble approach to combine the probabilistic forecasts from multiple teams provided in the form of quantiles. Each quantile of the ensemble forecast is constructed by taking the median of the forecasts across the models for the corresponding quantile.

Thus we note that there is a need for a comprehensive forecasting framework that combines multiple individual methods from various modeling paradigms within a performance-based ensemble. Instead of combining multiple contributed models, developing a single framework allows the models to share common datasets, runtime environments and improves overall robustness of the system. In our approach, we consider a combination of statistical, machine learning and mechanistic approaches, with a performance based ensembler to combine the models. Specifically, we use a variety of autoregressive methods (AR and ARIMA) [30] with exogenous variables, an Long Short-Term Memory (LSTM) model [14], nonlinear ensemble Kalman Filter (EnKF) [47], and compartmental SEIR model at the county level, and employ Bayesian Model Averaging (BMA) [16] to aggregate the individual forecasts. Further, since BMA computes the weighted average of probabilistic forecasts based on recent past performance, it allows us to provide robust forecasts while leveraging the best performing individual methods for different spatial regions and time points. Our approach is unique in the variety of methods incorporated for this task, and the only trained ensemble to the best of our knowledge for forecasting the COVID-19 pandemic in the United States.

Our key contributions and findings are as follows:

- We demonstrate the utility of a diverse, multi-model, performance based ensemble within a forecasting pipeline which can be quickly re-purposed for other pandemics and emerging outbreaks. Owing to the non-stationarity of the data, the individual methods and the BMA ensemble have to be trained periodically to keep pace with available disease surveillance. We thus describe a forecasting pipeline which has been operational for nearly 6 months, updated weekly at county level resolution.
- We highlight the challenges in building such an ensemble, and show how it leverages individual models at different time points and geographical regions. Through ablation analyses we show that at individual county level, some methods may be highly preferred, and contribute significantly to the ensemble.
- We compare the performance of our approach with our publicly available contributed models and *The Hub*’s unweighted ensemble using probabilistic scoring functions, and show that our model is ranked among the top three and increasingly preferred for longer forecast horizons.
- The framework we have described, in addition to being part of *The Hub*’s ensemble, also provides additional 1-2 weeks of synthetic data (i.e., forecast) for the mechanistic models to train on and produce more realistic projections for policy makers. One such model is publicly deployed on the Virginia Department of Health COVID-19 Data Insights portal^4^.

### 1.1 Related Work

As discussed previously, most of the COVID-19 forecasting approaches have involved mechanistic models [1, 6], including multiple variants that are part of *The Hub*. Traditionally, statistical and other data-driven methods have shown to be effective in epidemic forecasting but also rely heavily on high-quality data. Autoregressive (AR) models have been employed extensively in forecasting epidemics such as ILI and Dengue [26, 30, 34, 41, 46] and have yielded quality forecasts with the incorporation of exogenous variables such as social media and syndromic surveillance data sources. For COVID-19, linear models have been largely restricted to forecasting case time series at the national level and typically employ time series data from multiple regions to better model the data [5, 13, 22]. Modelers have also considered more complex systems such as deep-learning models. Specifically, Long Short-Term Memory (LSTM) networks owing to their ability to capture long-distance dependencies in a time series have been employed in ILI forecasting [36, 38, 40] with the inclusion of auxiliary data such as weather and geographical proximity casual model simulations. Another, model has been Graph Neural Networks which provide a natural framework to capture spatio-temporal interdependencies in epidemics dynamics [7, 42]. Some early examples of such models include [29] and [33] which incorporated auxiliary data and has been one of the long-standing national and state-level models in the *The Hub*. With the progression of the pandemic multiple papers have emerged [9, 20] that incorporate mobility data into GNNs to better model interventions. Recently, Gao et al. [11] proposed an attention-based GNN to integrate causal theory equations to regularize predictions. Finally, Bayesian model averagin (BMA) is a well-studied, effective framework for model averaging that, unlike the model selection, also takes into account the uncertainty in predictions. Its application to combining multiple weather models has been studied exhaustively by Raftery *et al* [27, 28], while its effectiveness in weighting competing ILI models has been demonstrated in [43, 44].

## 2 FORECASTING MODELS AND ENSEMBLE

Consider the COVID-19 confirmed cases time series 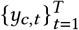 corresponding to a county *c* until *T*. The forecasting problem involves predicting forward *S*−steps ahead 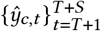 and we attempt to predict it by employing a variety of methods and then combining their forecasts. In the context of *The Hub*, we generate short-term forecasts of 1 to 4 week ahead, at weekly resolution. Since we are dealing with real-time systems and highly non-stationary data, the individual models as well as the ensemble model need to be retrained every week. As mentioned previously, each method consumes data in a different format and trains in slightly different ways. In this section, we briefly describe the individual models, associated data preparation and their training procedures.

### 2.1 Auto Regressive Models

This class of linear methods model the signal to be forecast using its lagged versions. We also incorporate the lagged time series data of other counties from that state to capture the spatial-temporal correlation between counties within a state. Since the time series under consideration is typically non-stationary, we log-transform it (to nullify the large variations in variance across time) and train the model every week over short segments with the assumption that the signal is relatively stationary over that period. The forecast model for county *i* is given by

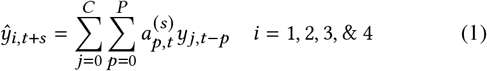

where *P* is the length of the training window. A sparse set of 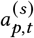 is estimated using an efficient bi-directional step-wise regression variable selection procedure [15, 30], a method that has proved effective in ILI and Dengue forecasting. Note that 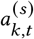 is time-varying and the model in (1) does not use rolling forecasts, that is, *ŷ*_*i,t s*_ is not incorporated in the forecast for *y*_*i,t s* 1_. This ensures that large errors do not propagate through the model (a trade-off as accurate forecasts are dropped). In the ensemble, we use two versions of this framework, the vanilla AR with only *i* = *j* and is denoted AR, while AR_spatial includes time series from other counties. After much experimentation, we set *P* = 7 for *AR* and *AR_spatial* with a training window of 8 weeks for both methods. The uncertainty in the forecasts were obtained by perturbing the coefficients by the training RMSE and run for multiple iterations to obtain the forecast distribution. Although we have experimented with other exogenous regressors such as aggregate mobility data, hospital visits, etc., they are not part of the current framework. The more general non-seasonal Autoregressive Integrated Moving Average (ARIMA) are effective for modeling signals with some degree of nonstationarity (trends), by specifying three parameters: autoregressive lag parameter *p*, the order of differencing *d*, and the order of the moving average filter *q*. We employ the popular forecast package [18] in order to determine the parameters. A separate model for each county is trained for each week and a training window of 10 weeks is considered. A range of parameters *p* [0, 3], *d* [0, 2], *q* [0, 3] was prespecified for the package to sweep through and the optimality of the model was determined using the Bayesian Information Criterion.

### 2.2 Long Short-Term Memory (LSTM) Networks

This method uses Long Short-Term Memory (LSTM) [14, 39] networks to capture the temporal dynamics of COVID-19 time series. Given the region’s time series X_*t*_ = x_*t T* 1_, …, x_*t*_, an LSTM model consists of k-stacked LSTM layers and each LSTM layer consists of *T* cells corresponding to *T* time points. The output of the last cell at the *k*th LSTM layer is fed into a fully connected layer to make the final prediction. Deep learning models usually require a large amount of training data which is not available in the context of COVID-19. Particularly, for counties of small population size where the surveillance data is sparse and noisy. Thus training a single model for each such region is highly susceptible to overfitting. In this respect, we explored a clustering-based approach that simultaneously learns COVID-19 dynamics from multiple regions within the cluster and infers a model per cluster. For the instance described in this framework, we grouped counties by their state, and trained one LSTM model per state. Other clutersing methods including k-means [12], time series k-means (tskmeans) [17] and k-shape [25] were also investigated but provide no significant improvement over geo-clustering method. The model was implemented as one LSTM layer with hidden size 32, one dense layer with hidden size 16 and a rectified linear unit (ReLU) [24] activation function, and one dropout layer with a dropout rate of 0.2. The output layer is a dense layer with linear activation and L2 kernel regularization with a 0.01 penalty factor. The historical window size is 3 weeks. We use the mean squared error (MSE) loss function and train the model with the Adam optimizer by setting a batch size of 32 and an epoch number of 200 using early stopping with a patience 50 for training. Probabilistic forecasts are generated using MCDropout [10].

### 2.3 Ensemble Kalman Filter

Ensemble Kalman Filter (EnKF) [4] is an approximate form of the Kalman filter that is particularly suited for practical applications. It represents the distribution of the state with sample mean and covariance which makes the filter updates computational less expensive compared to the update steps of the Kalman filter, and instead of a single model it takes an ensemble of model states that are propagated in parallel [45]. This is suited for nonlinear problems and large models since the ensemble covariance replaces the actual error co-variance matrix and avoids large computations. The ensemble mean is the best estimate for the actual state and the uncertainty of the forecast can be computed using the state covariance. The implementation of the Ensemble Kalman Filter [19, 23] involves three steps - initialization, prediction, and update. The ensemble members are randomly sampled around the estimate, and perturbations are added at each update and predict step. For model selection, we tried four different models using weekly and daily versions of incident and cumulative case series. The daily incident version was smoothed with a 7-day moving average to remove within week variations. Evaluation was done using median absolute percent error (APE) for counties withmore than 10 cases. The model with weekly cumulative case counts performed consistently well across different forecast duration. Using this model, we varied the number of samples from 5 to 2500 over forecasts generated retrospectively for a 25-week time period, with higher *N* leading to better performance at increasing computational costs. Based on the observed performance-speed trade-off, we set *N* = 100.

### 2.4 Compartmental SEIR Model

This method uses an Susceptible-Exposed-Infectious-Recovered (SEIR) model with outcome processing for case confirmation, hospitalization, and death, generated at a county-level resolution. While earlier variants of the model used inter-county travel and mixing (similar to [35]), due to time-varying social distancing and variable prevalence/incidence across counties, training a network model becomes difficult for an increasingly unsynchronized epidemic. Hence, in the model included in the ensemble, we train each county in isolation, where effects of social distancing and miscellaneous *adaptations* are captured as temporal variations in the SEIR model’s transmissibility term *β*. Using a simulation optimization approach, we sequentially estimate *β (t)*with appropriate delays and scaling applied from simulated infections to confirmed cases. For each *t*, the estimation is done using Golden Section Search (GSS), a maximally efficient extremum search method within a specified interval [21]. Each county’s confirmed cases is fit precisely through a daily varying transmissibility, and a smoothed version of recent *β* (*t*) is used for projections/forecasting. For this analysis we used the mean *β* (*t*) of recent 14 days, with the previous 21 days used for outlier removal (defined as a *β* (*t*) outside the 3σ range). We also perform 1-week ahead linear interpolation to smooth transitions for rapidly changing trajectories. While this method allows for layering counterfactual projections (i.e., increase or decrease in future *β*), we used the *status-quo* projection for the ensemble. Widespread pandemic eliminates sensitivity to initial conditions, hence we assumed steady low-level of importation/external seeding (1 case per 10 million). Other varied parameters include: duration of incubation (5-9 days) and infectiousness (2-7 days), case ascertainment rate (1x to 12x, depending on published seroprevalence testing), and delay from exposure to confirmation (4-12 days). The projected *β*’s for each cell in the experiment design are further randomized with 5 samples taken from the uniform distribution [0.8*β*, 1.1*β*].

### 2.5 Bayesian Model Averaging Ensemble

Since there is considerable variation in the case counts across counties, unlike [27, 44] we independently train a single BMA model per county. Considering *K* methods per county *M*_1_, *M*_2_, …, *M*_*k*_, we assume that the forecasts have a Normal distribution 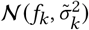. The BMA model assumes that the forecast *y* (note that we drop county *c* subscript as each county is trained independently) given the mean of the individual forecasts has a probability density

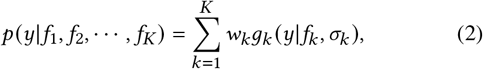

where *w*_*k*_ is the posterior probability of the *k*^th^ method’s forecast being the best one and is determined using *τ* training samples. (2) is a mixture of Gaussians and we proceed to determine the weights *w*_*k*_ and σ_*k*_. It is to be noted that despite each method providing its own uncertainty, we optimize further to refine it to obtain *σ*_*k*_. Given the distribution (2), the weights and variance parameters are obtained as the maximum likelihood estimate. The resulting log-likelihood function does not have an analytical solution, so we employ the standard expectation-maximization (EM) algorithm [2]. The optimization procedure is iterative and alternates between the E-step and the M-step with the updates for *w*_*k*_ and σ_*k*_ in the *j*^th^ iteration given by

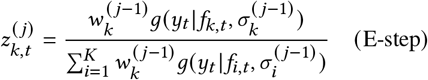

and

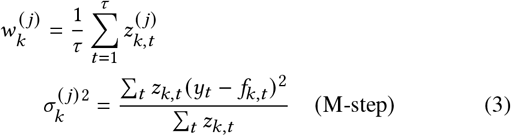

The EM steps typically converge to a local minima and the estimates are highly sensitive to initialization for which we consider 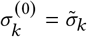 as a possible initial value.

## 3 THE PIPELINE

An overview of the forecasting pipeline is provided in Figure 1. As indicated, the individual models and the ensemble are updated weekly, using data until Saturday of the previous week. In order to align with the other models that are part of *The Hub*, we use confirmed cases at county level from the Johns Hopkins University Center for Systems Science and Engineering (JHU CSSE)[8] dashboard. Data updates are around 4AM EST, and each model consumes the 7-day smoothed version of confirmed cases data which are then preprocessed as described in Section 2. The AR, AR_spatial, LSTM and SEIR exploit parallelization and are run on multiple nodes of University of Virginia’s HPC cluster. ARIMA and EnKF are run on independent work stations. Simultaneously, the weights for BMA are also computed using the stored forecasts from previous weeks and the updated disease surveillance. Once the individual model runs are completed, the probabilistic forecasts for 1–to 4– week ahead are computed and formatted according to the guidelines. The forecasts are then checked to ensure that the forecast date and target dates are in agreement. We also verify if the forecast values for the individual models are below the county population. In the event it is over the population value the model forecasts for the particular location are disqualified and the model is rerun after inspection. If the individual model reruns are not complete by Sunday 8PM (ET), the model’s forecast is not incorporated into the week’s forecast and the BMA for that county is rerun with the respective method removed. Once the set of methods get approved, the forecasts are combined with the BMA weights to create probabilistic forecasts which are converted to quantiles and submitted to the GitHub repository of *The Hub* ^5^.

**Figure 1:**
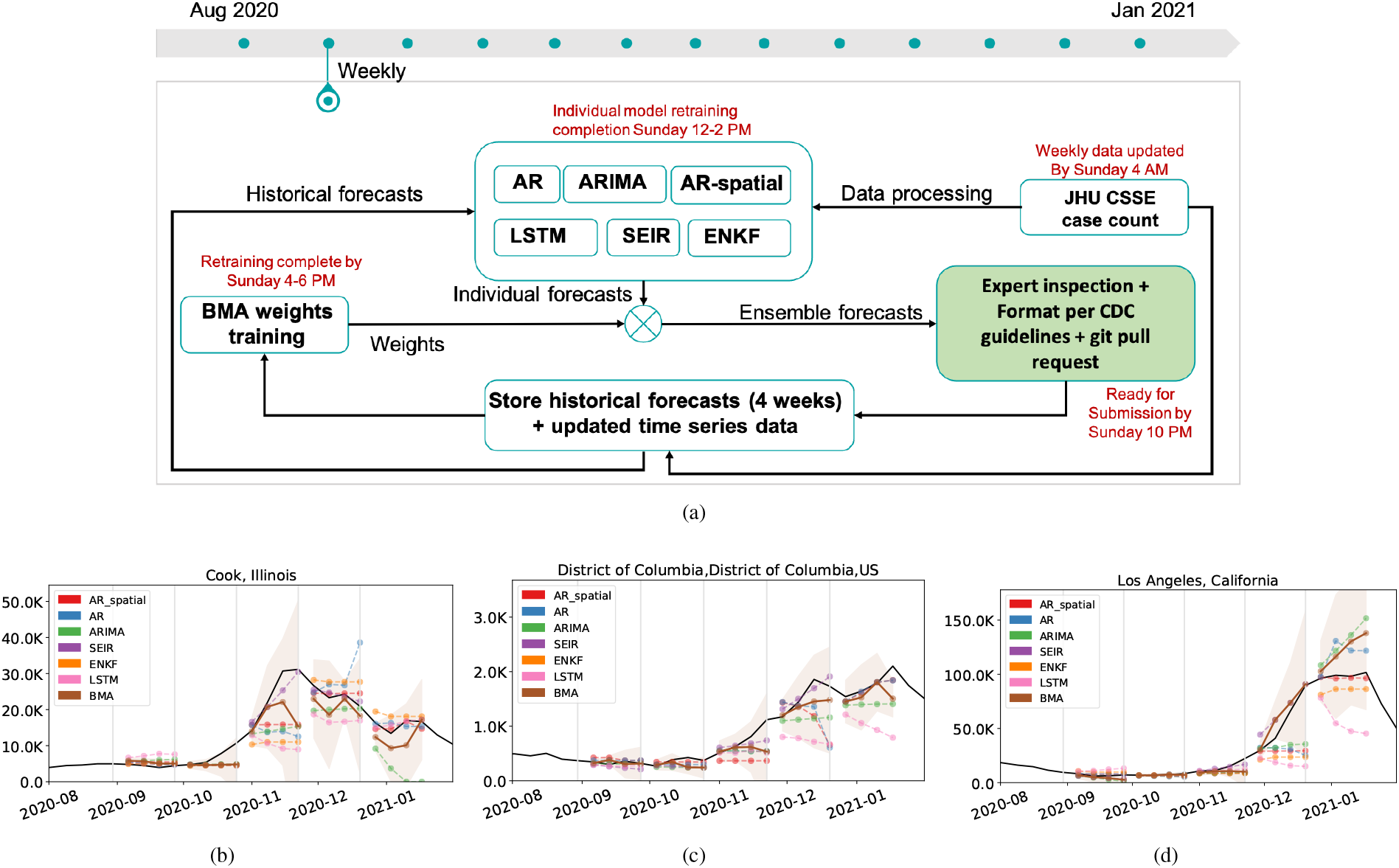
**Forecasting pipeline. (a) The weekly work flow, (b) Example forecasts for few counties with high case counts. Gray vertical lines indicate date of forecast (date of last observed data) and dashed lines to its right are the corresponding forecasts produced by the methods.**

In addition to being included in the *The Hub* ensemble, the BMA forecast is used to provide additional training data for the compartmental SEIR model. The SEIR model is useful for capturing potential future interventions and vaccination scenarios, and hence will benefit from a forecast that captures the statistical trends from individual models.

## 4 RESULTS

### 4.1 Forecasts and weights distribution

In an attempt to understand the spatio-temporal pattern of the dominant methods (the method with the highest weight) we present choropleth maps of US counties Figure 2(a) where each county’s color indicates the method with the highest weight for that particular week. In August 2020, we observe a nearly uniform distribution of weights. In November 2020, ARIMA appears to be dominating while in December 2020 the SEIR model is the dominant one. Figure 2(b) provides an insight into the number of counties a method is chosen for a particular week. Heat map suggests that ARIMA gets picked by many counties consistently while methods like EnKF dominate initially but in the later months LSTM starts to get higher weights for a lot more counties. Visual inspection indicates that each method dominates different regions at different times suggesting no spatial or temporal preferences for any particular method. We next provide a quantitative evaluation of the ensemble and its constituent methods using standard metrics.

**Figure 2:**
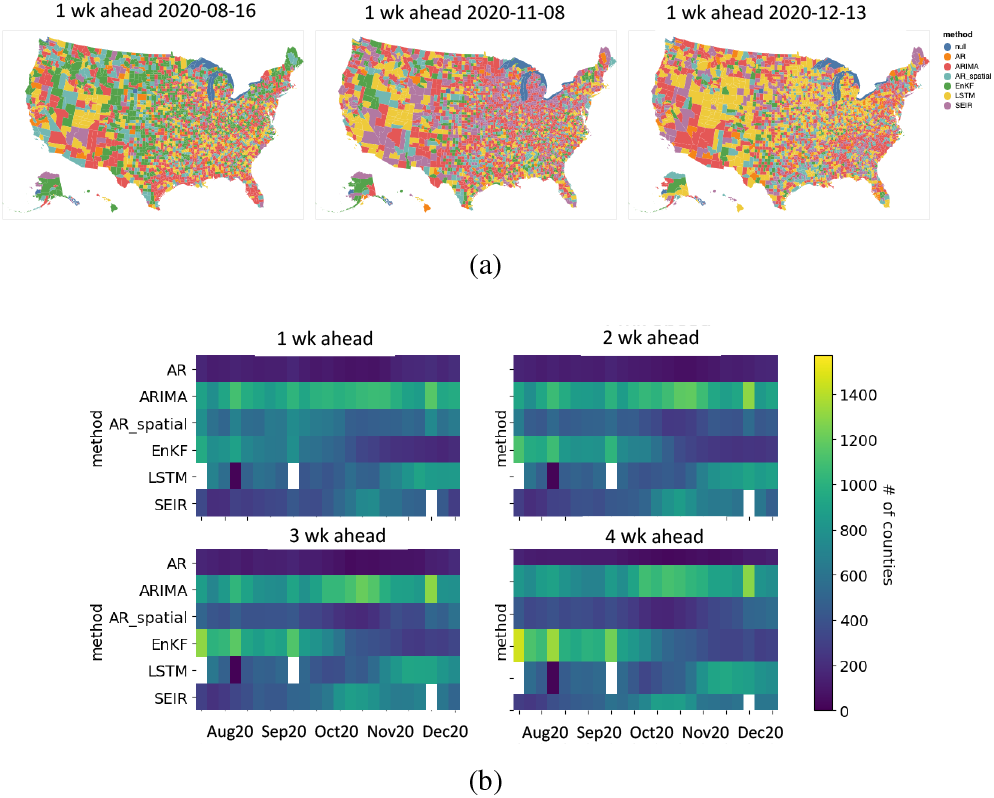
**Weights distribution: (a) SpAuag2t0iSaepl20dOicts20trNoivb20uDetci20on of methods with the highest weights for a county across three different forecast weeks. (b) A heatmap depicting the temporal evolution of the dominant methods across forecast weeks.**

### 4.2 A Comparison of Individual Methods

We evaluate 22 weeks of forecast data starting from the first week of August 2020 to the second week of January 2021 for all 3142 counties. The performance across 1-, 2-, 3-, and 4-week ahead targets (horizon) are evaluated separately. It is to be noted that since the county-level incident cases are typically small values. In all the evaluation, we consider only counties with observed forecast values greater than 10 cumulative cases. We employ mean absolute error (MAE) for comparing a set of *N* point forecasts 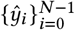 with the observed values 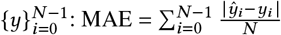

First, we consider the overall performance of each method by computing the MAE across all the forecast weeks and counties. The results are shown in Figure 3(a) and indicate that the average performance of individual models are similar. Also, the performance of BMA ensemble is comparable to, if not better than, the best performing model which is in accordance with previous observations made by [16, 43]. The average performance of the methods across all counties for each forecast week is shown plot Figure 3(b). In comparison to most methods the BMA has lower variations.

**Figure 3:**
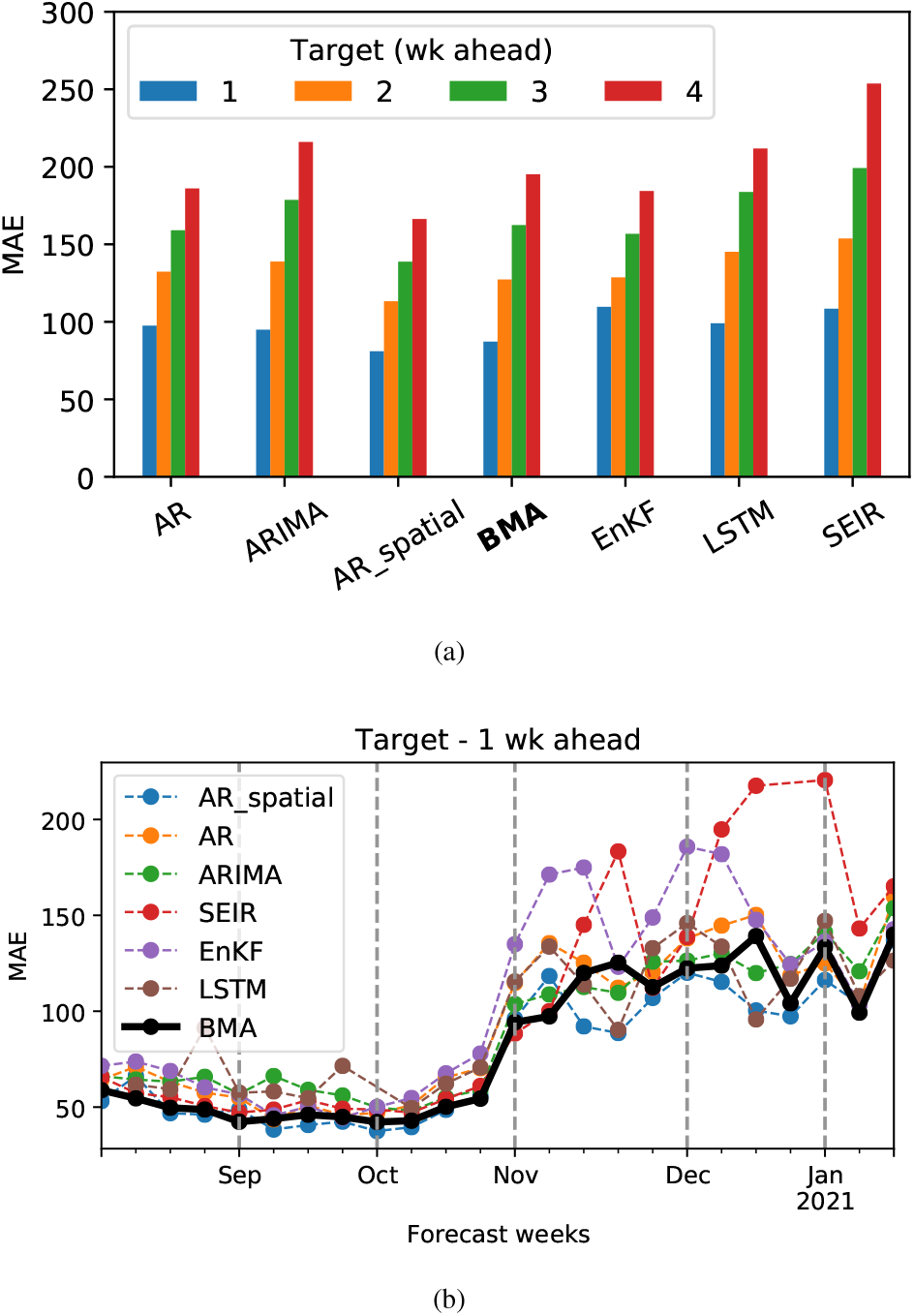
**The overall performance of individual methods and the BMA ensemble. (a) The overall performance is the MAE computed across all counties and forecast dates. The results indicate that average performance of BMA is comparable to method with the least MAE. (b) Performance of BMA and the individual models across various forecast weeks since August 2020. The performance of individual models vary across weeks and while the BMA shows relatively less variations across weeks.**

### 4.3 Relative Importance of Methods

To measure the relative importance of the models, we performed ablation analysis by omitting out forecast of a specific method in the ensemble. The ensemble is retrained and its performance is compared with the BMA trained using all models. In Figure 4(a) we show the results of the experiment. Although the drop in performance is significant it can be attributed to large errors incurred by making poor forecasts for counties with large number of cases. Further, this can be explained using Table 4(b) where we compute the change in MAE after the removal of a method. Let MAE^(*K*)^ denote the MAE for the BMA with all *K* models and let MAE^(*K*−1)^ denote the MAE of the BMA with a specific method removed. The % change in performance is computed as 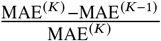. We picked some of the top COVID-19 hit counties and determined the best performing method across all weeks for that county. For Maricopa county, Arizona, SEIR model had the highest weights for eight weeks. When the SEIR model is dropped from the ensemble and the weights retrained, the relative change in performance is 1576.1%. This large change indicates that the other methods were unable to substitute for the SEIR model. These large errors are quite possible for individual statistical methods.

**Figure 4:**
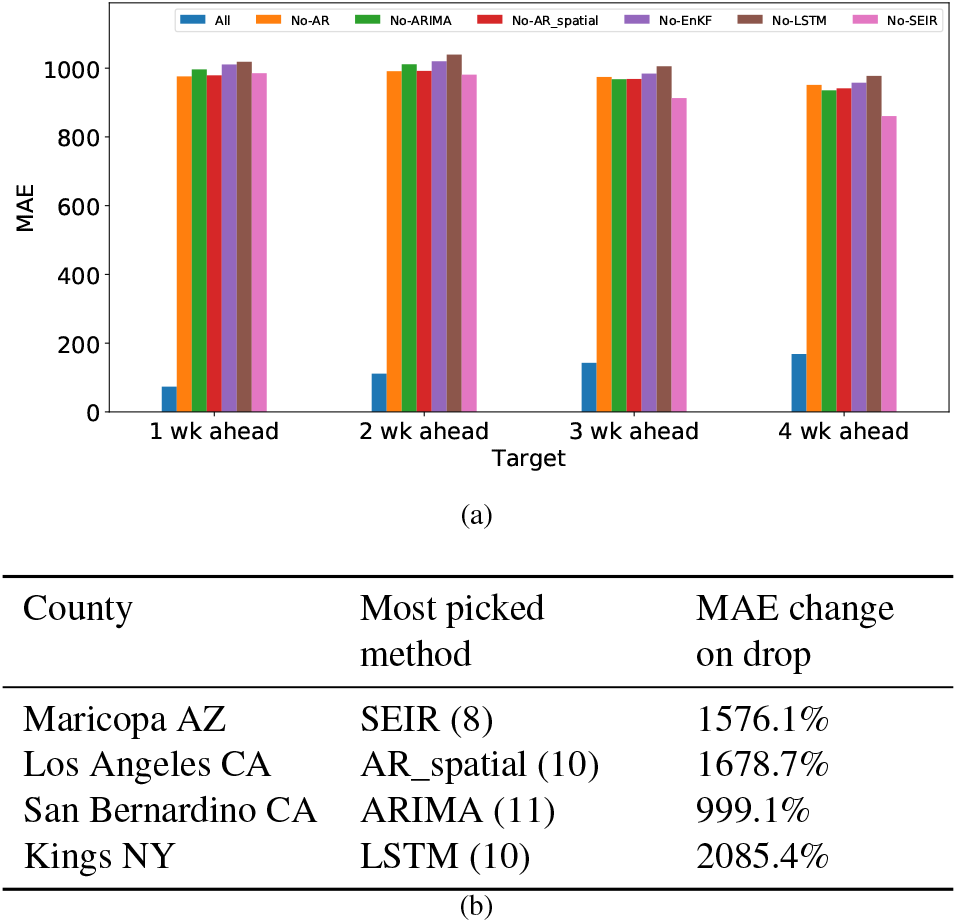
**Ablation analysis to study the relative importance of individual methods that feed into the BMA ensemble. The MAE is computed across all counties and forecast weeks. (a) A comparison of forecast performances of BMA^(*K*)^ with the other *K* BMA^(*K*−1)^ models. (b) % change in MAE for few county fore-casts after dropping the most picked method (computed across forecast weeks) with respect to the MAE of BMA with all methods included. The number of times a method is picked is indicated next to it.**

### 4.4 Comparison with *The Hub* Models

Among the 70 modeling teams present in *The Hub* only a handful of them provide county-level forecasts. In order to make a fair comparison, we only consider teams that have been providing consistent forecasts across most locations and targets since August 2020. The competing models are *COVIDhub-ensemble, JHU_IDD-CovidSP, LANL-GrowthRate*, and *CU-select* (the names are as specified in their respective submission to ForecastHub). Our forecasts are submitted under the team name *UVA_ensemble*.

JHU_IDD-CovidSP and CU-select are both county-level SEIR models that employ mobility to adjust for the disease spread parameters. LANL-GrowthRate, on the other hand, is a susceptible-infected model and determines the evolution of number of infections using a statistical model and then maps the infections to confirmed case data. The *COVIDhub-ensemble* uses ensembles forecasts from all eligible models (only models that produce all four week ahead forecasts) and takes the median prediction across all eligible models at each quantile. In order to make a comparison of probabilistic forecasts, we employ the interval score (IS) and its weighed version, an agreed upon metric in the community [3]. IS is defined as

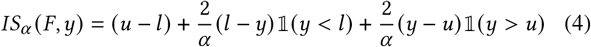

where (1 − *α*) × 100% is the prediction interval characterized by the upper bound *u* and the lower bound *l* that is likely to contain the forecast value *y*, and 𝕝 is the indicator function. The first term in (4) captures the spread while the second and the third term are penalties for under- and over-prediction, respectively. The IS is computed for various prediction intervals and their weighed combination yields the Weighted Interval Score (WIS):

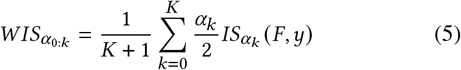

We compute the WIS for each modeling team’s forecasts and rank them accordingly for each target (horizon) and location. Rank 1 indicates the best performing team. In Figure 5(a) we plot the number of locations across the weeks that a model is ranked 1. We observe that across targets the UVA-ensemble is one of the top three performing models. We disaggregate the data into the respective months and plot the data in Figure 5(b). Mostly, COVIDHub-ensemble, CU-select, and UVA-ensemble are the top-performing models across most months but in December the performance of UVA-ensemble dropped. This drop in ranking could be due to multiple reasons; surges in cases and our model not being able to predict the trend; Model enhancements by other teams.

**Figure 5:**
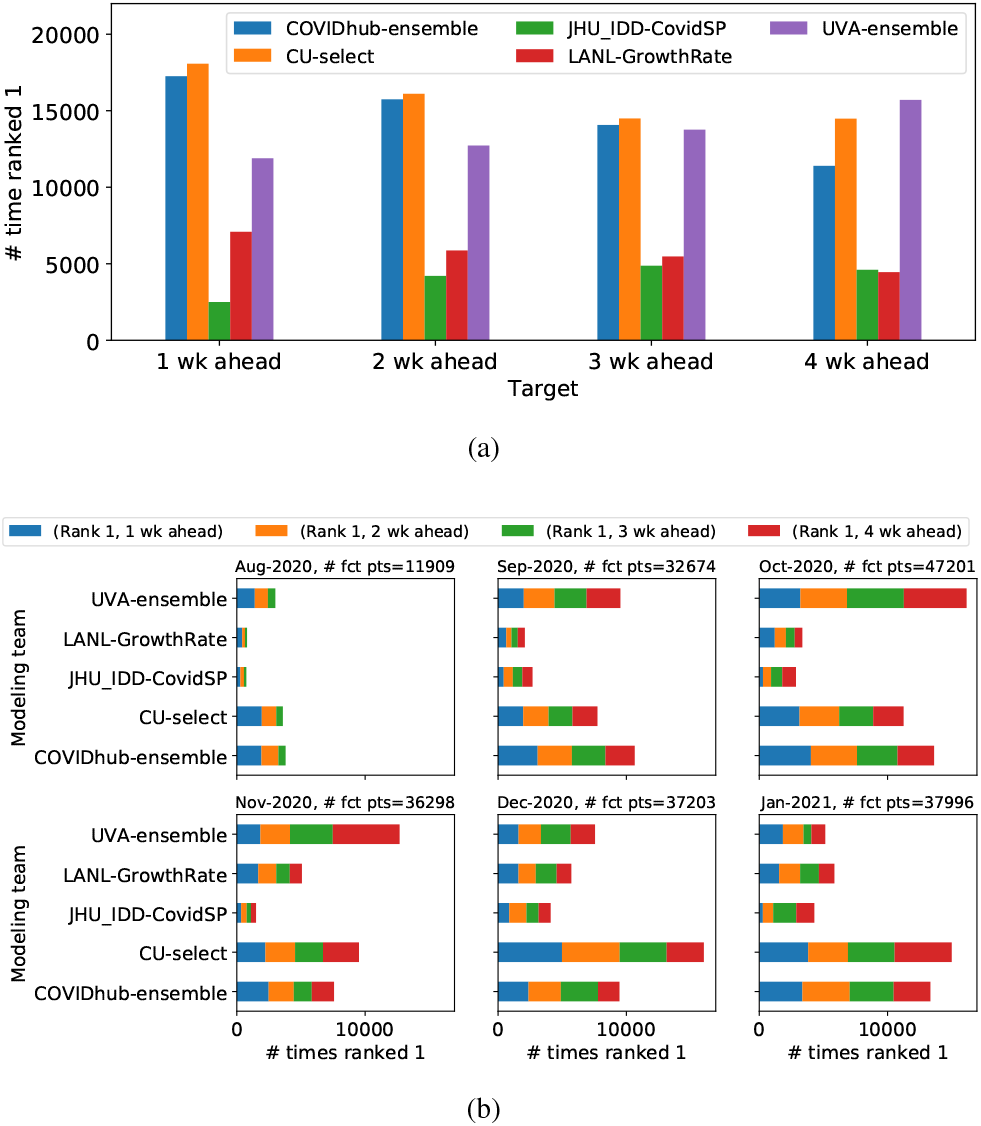
**Ranking of teams based on Mean WIS scores: (a) The plot indicates the number of locations across all weeks a given model was the best. (b) Performance of models across different months of 2020. The length of stacked bar indicates the total number of rank 1 assigned to a model while the individual stacks indicates the division for each target (horizon).**

## 5 DISCUSSION

Infectious disease forecasting is a rapidly evolving discipline with significant scope for improvement across tools, techniques, platforms and policy-making. As can be seen from the COVID-19 experience that, while simpler models are easy to setup, it is difficult to keep them regularly updated to ever-changing surveillance caused by epidemiological and socio-behavioral processes. Further, providing high resolution forecasts (e.g., at county level) require considerable scaling up and optimization of individual models to be able to provide regular updates. Understanding how the different data streams and modeling techniques can be integrated in a timely yet reliable fashion remains an open challenge. Depending on the epidemic outcome being forecast (e.g., cases, hospitalizations, deaths), they can be used to guide interventional measures, supplement healthcare resources or shape public messaging. While existing frameworks such as *The Hub* are quite successful in wrangling forecasts from multiple modeling teams for weekly updates to policymakers, our approach shows how a forecasting pipeline based on trained ensemble can still be beneficial under such dynamic conditions. While we have integrated methods from different modeling paradigm, given the available datasets and training approaches, an exhaustive search over the model space will still be challenging. Hierarchical model selection approaches along with ensembles that account for model complexity and diversity, will help produce more robust frameworks. Finally, using expert feedback as part of the loop, will help refine the objective functions by which the models are evaluated, thus providing more useful forecasts.

The codes related to the pipeline and an extended version of this paper containing more analysis can be accessed at https://github.com/aniruddhadiga/covid-19_forecast.

## Data Availability

Data and code will be made available upon request

## ACKNOWLEDGMENTS

The authors would like to thank members of the Biocomplexity COVID-19 Response Team and the Network Systems Science and Advanced Computing (NSSAC) Division for their thoughtful comments and suggestions related to epidemic modeling and response support. We thank members of the Biocomplexity Institute and Initiative, University of Virginia, for useful discussion and suggestions. This work was partially supported by National Institutes of Health (NIH) Grant 1R01GM109718, NSF BIG DATA Grant IIS-1633028, NSF Grant No.: OAC-1916805, NSF Expeditions in Computing Grant CCF-1918656, CCF-1917819, NSF RAPID CNS-2028004, NSF RAPID OAC-2027541, US Centers for Disease Control and Prevention 75D30119C05935, University of Virginia Strategic Investment Fund award number SIF160, and Defense Threat Reduction Agency (DTRA) under Contract No. HDTRA1-19-D-0007.

## Footnotes

1 https://covidtracking.com/

2 https://nssac.bii.virginia.edu/covid-19/dashboard/

3 https://viz.covid19forecasthub.org/

4 https://www.vdh.virginia.gov/coronavirus/covid-19-data-insights/

5 https://github.com/reichlab/covid19-forecast-hub

## Notes

### Competing Interest Statement

The authors have declared no competing interest.

### Funding Statement

The authors would like to thank members of the Network Systems Science and Advanced Computing (NSSAC) Division for interesting discussion and suggestions related to epidemic science and machine learning. This work was partially supported by National Institutes of Health (NIH) Grant 1R01GM109718, NSF BIG DATA Grant IIS-1633028, NSF DIBBS Grant ACI-1443054, DTRA subcontract/ARA S-D00189-15-TO-01-UVA, NSF Grant No.: OAC-1916805, US Centers for Disease Control and Prevention 75D30119C05935 and a collaborative seed grant from the UVA Global Infectious Disease Institute.

### Author Declarations

This research work includes all publicly available data and does not use patient data

